# Spatial Network based model forecasting transmission and control of COVID-19

**DOI:** 10.1101/2020.05.06.20092858

**Authors:** Natasha Sharma, Atul Kumar Verma, Arvind Kumar Gupta

## Abstract

The SARS-CoV-2 driven infectious novel coronavirus disease (COVID-19) has been declared a pandemic by virtue of its brutal impact on the world in terms of loss on human life, health, economy, and other crucial resources. With the aim to explore more about its aspects, we adopted the *SEIQRD* (Susceptible-Exposed-Infected-Quarantine-Recovered-Death) pandemic spread with a time delay on the heterogeneous population and geography in this work. Focusing on the spatial heterogeneity, the entire population of interest in a region is divided into small distinct geographical sub regions, which interact by means of migration networks across boundaries. Utilizing the estimations of the time delay differential equations based model, we analyzed the spread dynamics of disease in a region and its sub regions. The model based numerical outcomes are validated from real time available data for India. We computed the approximate peak infection in forward time and relative timespan when disease outspread halts. To further evaluate the influence of the delay on the long term system dynamics, the sensitivity analysis is performed on time delay. The most crucial parameter, basic reproduction number *R*_0_ and its time-dependent generalization, has been estimated at both regional and sub regional levels. The impact of the most significant lockdown measure that has been implemented in India to contain the pandemic spread has been extensively studied by considering no lockdown scenario. A suggestion based on outcomes, for a bit relaxed lockdown, followed by an extended period of strict social distancing as one of the most effective control measures to manage COVID-19 spread is provided for India.

## I. INTRODUCTION

The novel coronavirus disease, officially known as 2019-nCoV, or SARS-CoV-2 (severe acute respiratory syndrome coronavirus 2), commonly called COVID-19, has provided the world with an unparallel challenge. The first reported case in the COVID-19 outbreak appeared in the Wuhan city of China in December 2019, and since then, it has rapidly spread in 210 countries and territories around the world [1]. As of April 30, 2020, a total of 3,275,475 confirmed cases of the coronavirus and a toll of 231,576 deaths has been reported globally [2]. At this moment of time, there is no definite vaccine or treatment for COVID-19. It has thrown an unpredictable burden on healthcare systems in the majority of the developed as well as developing countries. With its breakneck spread all across, World Health Organization(WHO) has declared it as a pandemic and to contain the upsurge by breaking its person to person transmission chain involving significant human migration, major steps including partial or complete lockdowns, international travel bans, and social distancing has been widely adopted.

In India, the first case of coronavirus pandemic was identified on January 30, 2020, originating from China. Since this appearance, there has been a continuous rise in the number of infections with the total number of 33,610 cases on April 30, 2020, of which there are 8,373 recoveries and 1,075 deaths [3]. As a preventive measure, India also implemented a strict lockdown along with curfew in certain states, limiting movement of the entire population to minimize social contacts. The efficiency of such measures is geographically sensitive due to different population densities, social contact networks, and health care facilities. Though these measures are important for controlling the virus, they have an extensive burden on the global economy as well. Quantitative projection of the effect of these measures in reducing infection is crucial in outlining social and economic policies. Further, understanding the transmission dynamics of COVID-19 not only provides deep insights into the epidemiological situation to enhance public-health planning, but such investigations can also aid in the layout of alternative outbreak control measures.

Knowledge of the early spread dynamics of the infection and figuring out the capability and performance of applied control measures is critical for assessing the extensive outspread to occur in new regions. Besides medical and biological research, understanding the urgency to develop a predictable mathematical model for the COVID-19 outbreak, few mathematical studies based on statistical reasoning and simulations have been taken up in past months [4–6]. Later, accepting the challenges to explore the efficiency of various control measures since the outbreak, fewer studies adopted more appropriate dynamical equations based on mathematical modeling techniques. Compartment models such as *SI*, *SIR, SEIR* which exhibits the change in the category of population among the susceptible (*S*), exposed (*E*), infected (*I*) and recovered (*R*) classes have been developed and studied to understand the spread pattern of COVID-19 in many countries [7–11].

Further, as reported, COVID-19 has a latent/incubation period (time from exposure to the development of symptoms), which is estimated to be between 2 and 14 days [12]. This incubation period is very significant as it allows the health authorities to introduce more adequate systems for separating people suspected of carrying the virus, as a way of controlling and preventing the spread of the pandemic [13]. The inclusion of the latent period in compartment models gives rise to COVID-19 time delay models in which the dynamic behavior of the disease at time *t* depends on the state of the system at a time prior to the delay period [14, 15]. However, these studies assume homogeneity in a large population and ignore numerical variations originating from natural births, deaths, and human migration networks across the regions. To the amount that population and geographic heterogeneities play crucial roles in the infection outbreak process, in order to pick up the vital characteristics of the pandemic, it is advantageous to include them in the time delay model.

Capturing the critical impact of the delay period of COVID-19, this study presents a mathematical model to analyze the spread of the novel coronavirus within a heterogeneous population and geography. We propose a more realistic time delay differential equations based *SEIQRD* model by incorporating quarantined (*Q*)- confirmed and separated infected population as well as death due to infection class (*D*) along with the optimal values for model parameters, which reasonably fit the actual infection cases of the pandemic. Spatial and stochastic parameters like heterogeneous population, natural births, deaths, and population migration networks within a geographical state (subpopulation in a country) boundaries have been considered. Predominantly, in the presented model, considering the spatial heterogeneity, the entire population of the broad geographical region of interest is divided into small sub regions-states, which interact by means of migrations across the state boundaries. Moreover, there are many inevitable questions related to the spread of the pandemic. How many people are at risk of infection? When will be the infection at its peak and when will it end? Are the existing control measures sufficient to control the outbreak? With the aim to explore more about the transmission of COVID-19 dynamics and to predict its potential tendency, the COVID-19 pandemic is estimated for forward time. The general points of interest including total infected cases, infection peak, pandemic ending time are investigated. The most vital parameter, basic reproduction number *R*_0_ and its time-dependent generalization that decides if an originating infectious disease can extend in a population, has been estimated at both regional and its sub regional levels. Additionally, the influence of the time delay on the long term pandemic dynamics has been explored. Further, we use the model to examine the impact of the most significant lockdown measure that has been implemented to contain the pandemic spread.

## II. MATERIAL AND METHOD

### A. Generalized *SEIQRD* model with time delay

To estimate the trajectories of COVID-19 transmission, in the proposed approach, a vast geographic region with heterogeneous population distribution (country) is partitioned into *n* smaller sub-regions (states). To take into account the effect of both inter and intrastate infectious population, migration for these states are explicitly incorporated (Fig. 1(a)), using population migration adjacency matrix *λ* (with order *n* × *n*). The diagonal coefficients of this matrix represent the migration rate within a state while for any of its *j^th^* row, *k^th^* element indicates the migration factor from the *j^th^* state to the *k^th^* state. For any two states which share no boundaries migration rate is 0. Within a given state, the transmission is handled according to a deterministic compartmental model dividing the population into six epidemiological classes *S*(*t*), *E*(*t*), *I*(*t*), *Q*(*t*), *R*(*t*), *D*(*t*) describing at time t the respective proportion of the susceptible cases, exposed cases (infected but not capable of transmitting infection, in a latent period), infected cases (with the infection spreading capacity and not yet separated from those who are not infected), quarantined cases (confirmed and separated infected), recovered cases and deaths owing to virus cases. Fig. 1(b) shows the movement within the classes in model with time delay. Since the latent period (*τ*) of the COVID-19 is as long as 2 to 14 days [1], the exposed class (*E*) originates because of interactions with the amount *β* within susceptible (*S*) and infected class (*I*). Only those members of the exposed class who survive the latency period move to the infected class with a rate of *σ*. The infected population tested positive for COVID-19 moves to quarantine class (*Q*) for further treatment with parameter *ϵ*. After a course of quarantine treatment, either a part of its population is discharged with the rate *δ* from the hospital (*R*) or encounter death due to underline diseases (*D*) with an estimate *γ*. An appendix (*A*) provides complete details of our mathematical model characterized by a group of ordinary differential equations depicting the outbreak at time *t*. The primary model parameters, reasonably fitting the actual infected cases till date, are described in Table I in the appendix (*C*).

**Figure 1:**
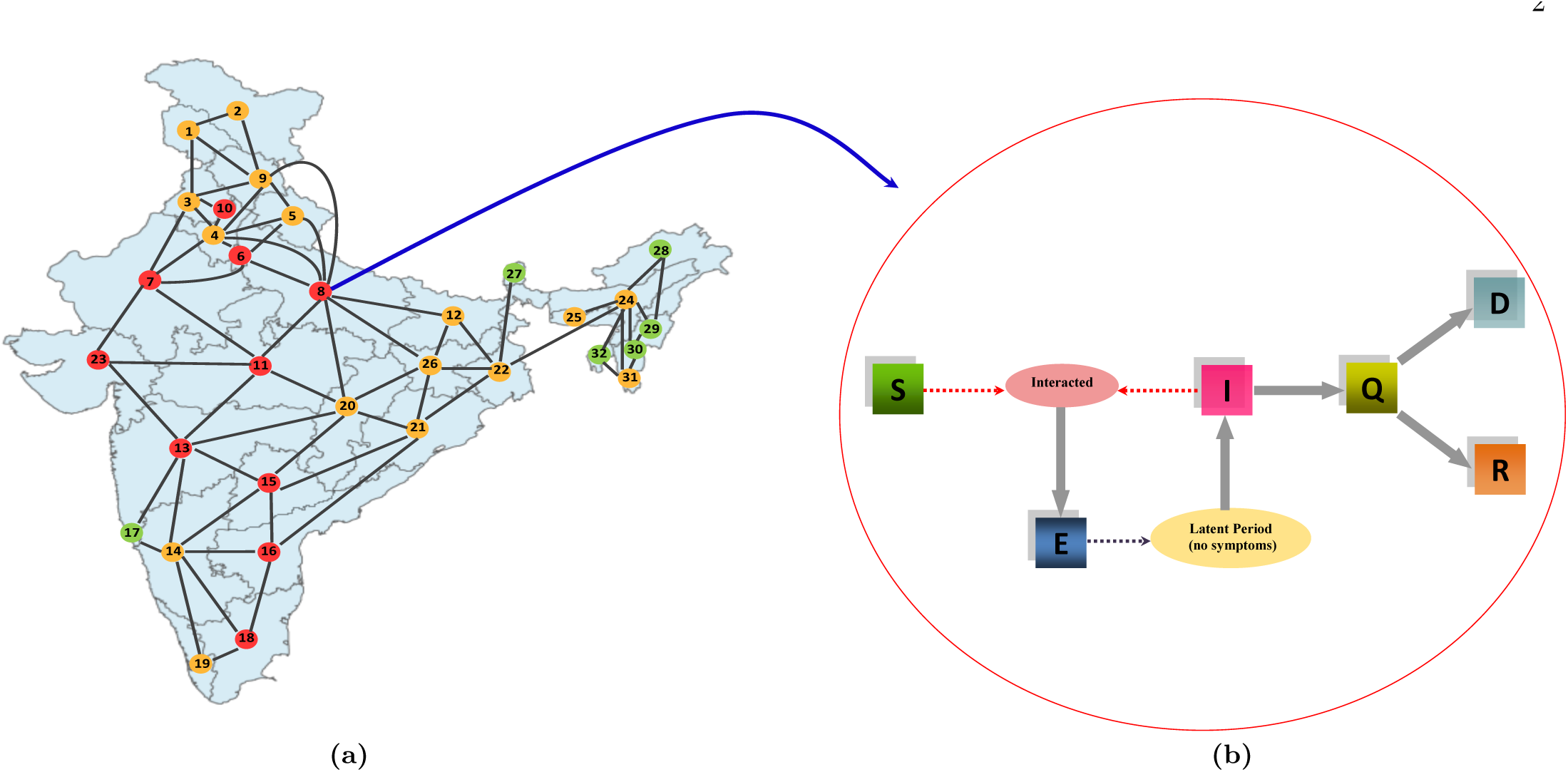
(a) The map of India along with different states (represented by filled dots, numbered from 1 to 32^*^). The solid lines shows the connections of a state with its neighboring states resulting in inter state movements. The states with more/less than 2000 cases as on April 30, 2020 are shown by red/orange color. COVID-19 free states are represented by green color.The complete dataset reflecting state wise population distribution is provided in Appendix (*C*) in Table II.^*^ Note that in the study we have not considered a few Indian states and Union Territories including Lakshadweep, Dadra and Nagar Haveli, Daman and Diu, Andaman and Nicobar Islands and Puducherry on account of their comparatively low population density. (b) Within a state, Schematic of *SEIQRD* Model with time delay.

**Table I:** Quantification of parameters value used in the proposed model.

| Parameter | Value | Source (Ref.) |
| --- | --- | --- |
| $\sigma$ | 1/7 | [1, 2] |
| $\delta$ | 0.22 | [18] |
| $\gamma$ | 0.034 | [1, 2] |
| $\tau$ | 3-14 | [1, 2] |
| $\beta$ | 1-13 | [18] |
| Entries of $\lambda$ | 0.1 – 0.8 | [24] |

### B. Data Source

The epidemic bulletin from the World Health Organization provides us with primary data on epidemiological research. According to the reports by WHO, the incubation period for COVID-19, which is the time between exposure to the virus (becoming infected) and symptom onset, is, on average 7 days; however, it can be up to 14 days [1]. The average mortality rate based on historical data is about 3.4% [2, 11].

To check how optimal is the fitting between the actual official data for the pandemic spread and the model based obtained numerical approximations, we use the statistical Kolmogorov-Smirnov (K-S) test [16, 17]. The K-S goodness-of-fit test is performed on the data to ascertain whether two data series are from the same continuous distribution or not. The null hypothesis implies that data samples are from the same continuous distribution, and the alternative hypothesis states the opposite. The statistic *h* is 1 if the test rejects the null hypothesis at the 5% level of significance, and 0 otherwise. For the proposed model with retained parameters, the results indicate (with *h* = 0) that both the actual and the projected number of cases are from the same continuous distribution, as desired, showing optimistic estimations.

## III. RESULTS

In this section, we conduct a more detailed model analysis to depict outspread for the COVID-19 pandemic. We obtained the numerical solution of the mathematical model discussed in the appendix (*A*) based on the chosen set of parameters for India. Using suitably preferred parameters, the same detailing can be obtained for any geographic region. Moving further, we aim at forecasting the infection cases along with the basic reproduction number. Moreover, we use the model to examine the impact of the most significant lockdown measure that has been implemented in India to contain the pandemic. It is significant to mention that, at present, due to lack of sufficient diagnostic test for COVID-19, the total number of infected cases (I + Q) in any *j*^th^ state, can be given by (*I*(*t*) + *Q*(*t*))*αN_j_*. Here, *N_j_* denotes the population of the *j^th^* state, and *α* is considered approximately as 0.1 in the role of the current testing events [18]. Clearly, the total infected cases in a country with n states can be obtained by the sum of corresponding data from the states.

At the onset, we used the proposed model to fit actual available COVID-19 infection cases for India up to April 30, 2020 (Fig. 2). Notably, since India went into a strict lockdown that commenced on March 25, 2020, officially published data by Ministry of Health and Family Welfare (MoHFW), Government of India from March 16, 2020 to April 30, 2020 are marked in pink spots and are considered as a direct validation source. As expected, we can clearly see that the prediction of the exact number of cases diagnosed in the past time by the model is in reasonable agreement with the real value. With an aim to estimate the potential tendency of the COVID-19 pandemic and to examine the impact of the lockdown control measure we compared the infection spread in the cases of current strict lockdown and no lockdown in place as under:

**Figure 2:**
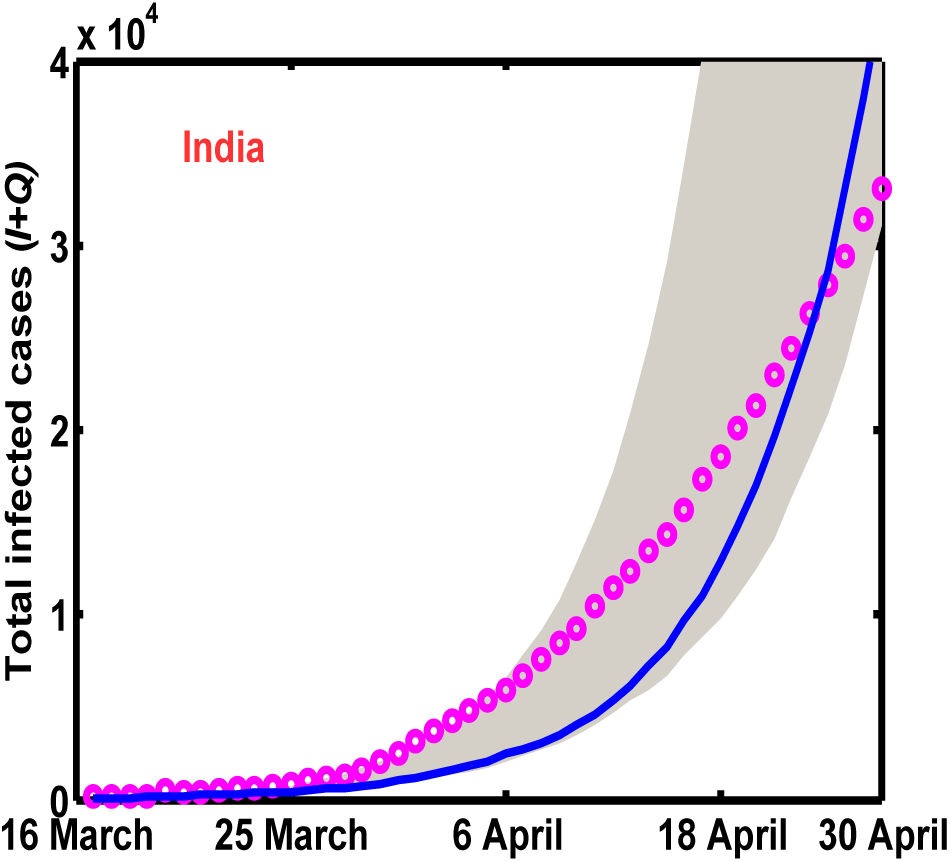
(Color Online) Fitting of the actual available total COVID-19 infected cases (*I* + *Q*) shown in pink colored markers for India from 16 March to 30 April, 2020. Solid blue line represents the fitted curve with parameters: intrastate population migration factor = 0.34, interstate population migration factor = 0, *β* = 3.319, *τ* = 8.8, *μ* = 0.0412, *δ* = 0.22, *σ* = 1/7, *ϵ* = 0.85, and *γ* = 0.034. The shaded region is the bound for the fitted model with respect to *τ* ∈ [6, 12].

***Nationwide strict Lockdown:*** With parameters in hand, we executed the model forward in time to visualize the progress of the pandemic in the present situation of strict lockdown. Fig. 3 shows the trajectory for the total number of infected cases in India (Infectious + Quarantined) in the coming months. Fig. 3(a) displays a five month forecast in the present scenario of nationwide lockdown as a strategic move to control the pandemic spread. With strict lockdown measures currently in place, since all transport services: road, air, and rail were suspended, we set the interstate population migration index to null in the population migration adjacency matrix *λ*, and the average contacts between susceptible and infected are considered to be proximately three. Based on these criteria, our model featured a single infection peak on June 30, 2020 with the total number of estimated infected to be around 5.2 million. The outbreak is expected to be nearing its end by late August, 2020.

**Figure 3:**
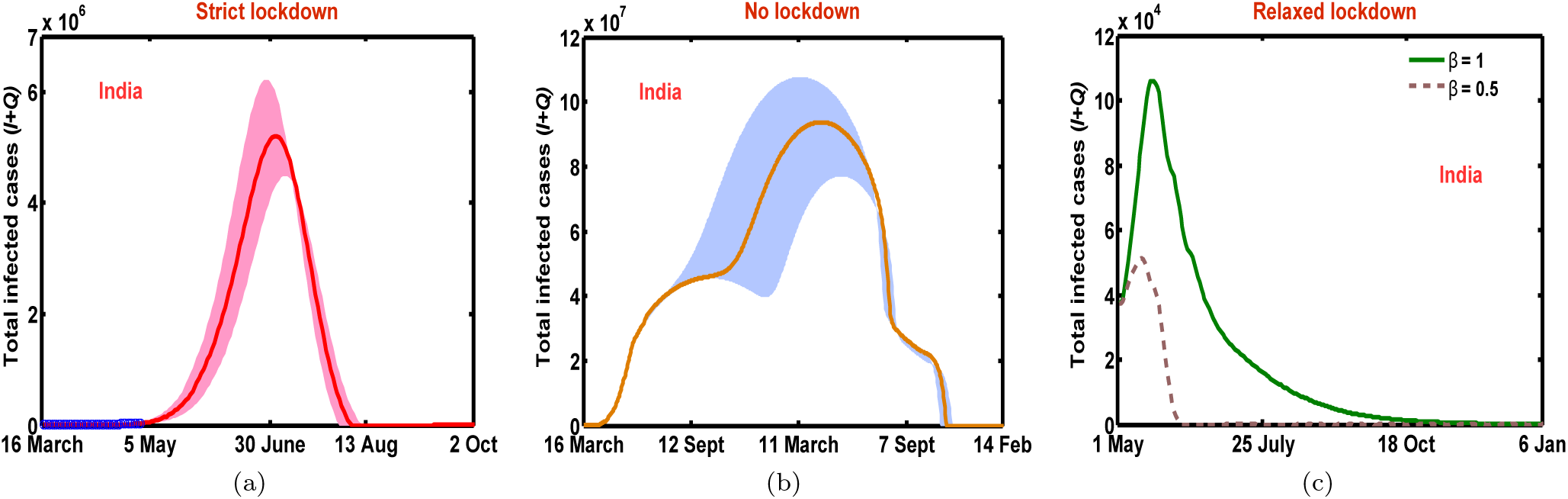
(Color online) Total infected cases (*I* + *Q*) with respect to time (in days) in different scenarios (a) strict lockdown (b) no lockdown (c) relaxed lockdown. In (a) blue line represents real data from 16 March to 30 April, 2020. The parameters are (intrastate population migration factor, interstate population migration factor, *β*) = (0.34, 0, 3.319), (0.8, 0.6, 9), (0.34, 0.10, 0.5,1) in (a-c) with common parameters *τ* = 8.8, *μ* = 0.0412, *δ* = 0.22, σ = 1/7, *ϵ* = 0.85, and *γ* = 0.034. The shaded region in (a-b) is the bound for the fitted model with respect to *τ* ∈ [6, 12]. In (b) the dates 11 March, 7 Sept are for year 2021, 14 Feb for 2022; in (c) 6 Jan belong to year 2021; while remaining dates are from 2020.

***No Lockdown:*** To highlight the effectiveness of the ongoing national lockdown, in comparison, Fig. 3(b) shows infection forecast if in case lockdown was not implemented. In this situation, we allowed normal migration within the states and across the boundaries by setting non zero estimates for diagonal and off diagonal entries in the migration adjacency matrix. However, assuming that some form of control measure would continue to be in the system to reduce social contact, we choose average contacts between susceptible and infected to be approximately nine [18]. As anticipated, if interventions would not have been there, India might have experienced 18 times worst the numbers with 40 million infections by mid of October, 2020 with a peak of approximately 90 million cases around April, 2021. There still would have been around 5, 00, 000 infectious cases revealed at the end of August, 2021 as exhibited by Fig. 4(b).

**Figure 4:**
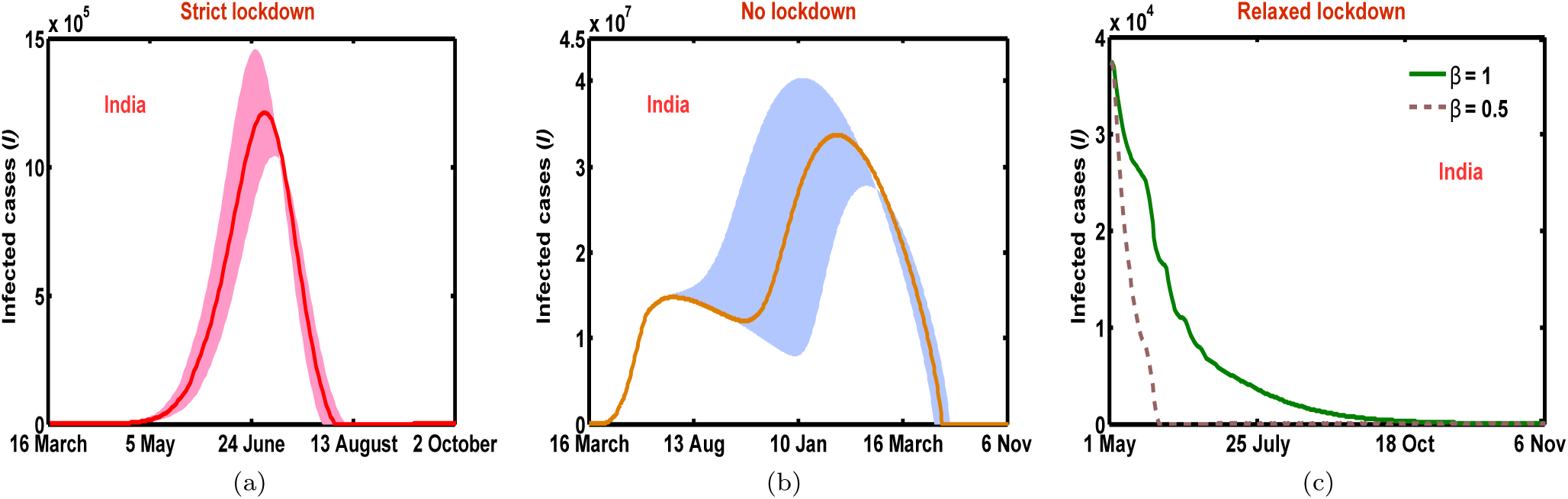
(Color online) Infectious cases (*I*) with respect to time (in days) in different cases (a) strict lockdown (b) no lockdown (c) relaxed lockdown. The parameters are (intrastate population migration factor, interstate population migration factor, *β*) = (0.34, 0, 3.319), (0.8, 0.6, 9), (0.34, 0.10, 0.5,1) in (a-c) with common parameters *τ* = 8.8, *μ* = 0.0412, *δ* = 0.22, *σ* = 1/7, *ϵ* = 0.85, and *γ* = 0.034. The shaded region in (a-b) is the bound for the fitted model with respect to *τ* ∈ [6,12]. In (b) the dates 10 Jan, 16 March, 6 Nov belong to year 2021 while remaining dates are from current year 2020.

***Proposed Scenario:*** In the third scenario, Fig. 3(c) panel exhibits a rundown for a proposed protocol with a bit relaxed lockdown, allowing restricted movements within and across the state boundaries beyond May 3, 2020. This relaxation assumes gradual freedom of movement for essential activities only, thereby ensuring the upliftment of lockdown in stages to save the economy. Moreover, considering self awareness within the population, we invoked reduced average contact parameter ft between susceptible and infected to be approximately one, through social distancing impacts in this case. This alteration was required as a step to break or lessen human to human transmission chain of the virus. As likely, this consideration brings the total number of infections to comparatively lower values and slows the spread of the disease to manageable levels. Moreover, peak infections in the proposed scenario decrease significantly in comparison to a strict lockdown situation. This observation clearly highlights the fact that social distancing is one of several changes that we have to adapt strictly in the coming weeks. To ensure the distances are well maintained at work or public places, more mature ways of monitoring, including artificial intelligence or machine learning software, can be used. As depicted in Fig. 3(c), as a suggestion, if social distancing is enforced broadly and is maintained in the coming months, the number of new infections would decrease to a significantly submissive level, and the outbreak could eventually be controlled.

Additionally, in order to further evaluate the influence of the delay on the long term disease dynamics, we performed the sensitivity analysis on *τ* keeping other parameters fixed. We plotted the total infected cases and infectious cases as a function of time for systematically varied values of *τ* ∈ [6,12] shown by shaded regions in Fig. 2, Fig. 3 and Fig. 4. Apparently, a small change in delay has no qualitative impact on the nature of the spread, apart from shifting of the curve, thereby preserving the basic shape of the infection pattern. Shifts in infection peak with varied *τ* provide us with a possible domain window range for estimation of infection cases owing to regional differences in India on account of health, education, religion, and per capita income.

In order to capture *R*_0_ for heterogeneous population distribution in India, we computed its value using the proposed model by the relation given by Eq.(7) in the appendix (*B*). The estimated basic reproduction number for India in the present scenario of nationwide lockdown turns out to be 1.18. Based on this value of *R*_0_, as expected, from the transmission dynamics of the infection in a strick lockdown situation, shown in Fig. 4(a), the number first increases before tending to zero, thereby *R*_0_ = 1 acts as a clear edge between the disease widely spreading or dying out. It is worth mentioning here that in case of a relaxed lockdown with average contact rate between susceptible and infected as one, *R*_0_ turns out to be 0.8739, which is much desired for outspread levels to reduce further to a manageable extent. This can be clearly verified from Fig. 4(c), where the number of infectious decreases monotonically to zero. Additionally, Fig. 5 shows the time-dependent effective basic reproductive number 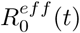 corresponding to the infection trajectories in Fig. 4(a). As hoped for, this value is greater than unity before peak infection and smaller than unity beyond its peak, serving as a useful estimate of the local rate of change of infectives at any time. It is noteworthy to mention here that for any state of India, R_0_ can be obtained based on the relation given by Eq.(8) in the appendix (*B*).

**Figure 5:**
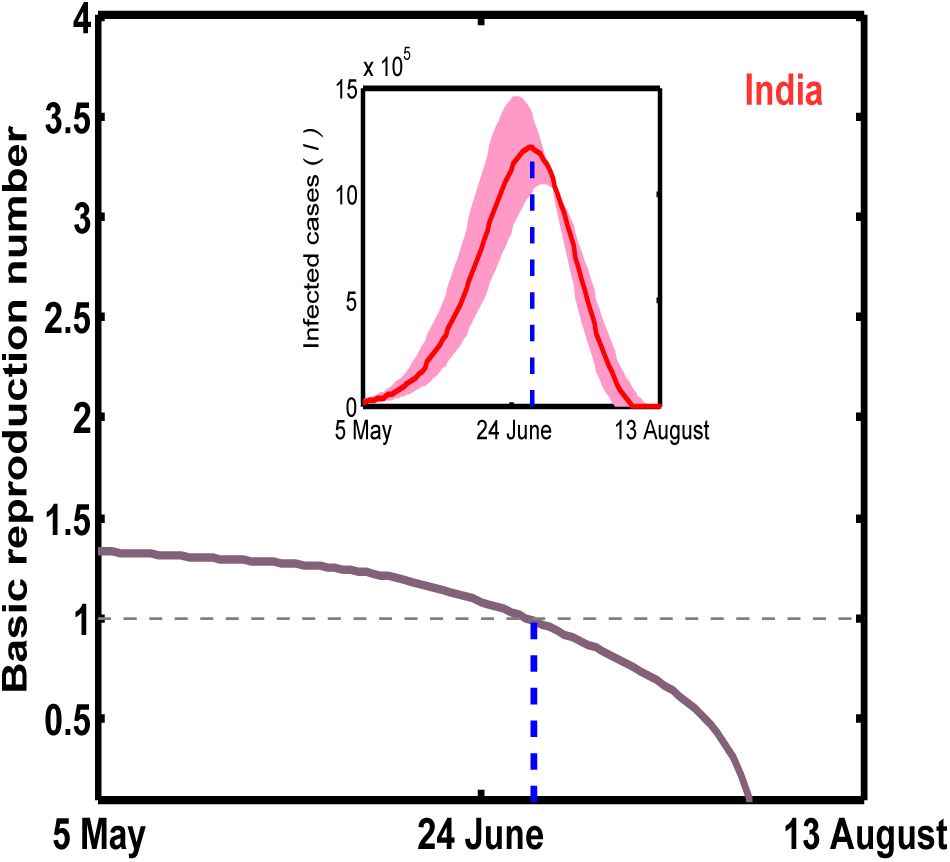
(Color Online) Basic reproduction number with respect to time in presence of strict lockdown for intrastate population migration factor = 0.34, interstate population migration factor = 0, *β* = 3.319, *τ* = 8.8, *μ* = 0.0412, *δ* = 0.22, *σ* =1/7, *ϵ* = 0.85, and *γ* = 0.034. The inset figure shows infectious cases as in Fig. 4(a) with time for same parameters.

The model discussed here, could be helpful to health authorities for depicting the total number of infection cases along with the peak infection. The estimated fit values could be made better on a daily base as more data become available.

## IV. DISCUSSION

We have presented a generalized *SEIQRD* mathematical model with a time delay to analyze the spread of COVID-19 infection in a population. The proposed model incorporates spatial and stochastic parameters like heterogeneous population, natural births, deaths, and population migration networks within geographical state boundaries. Based on a detailed analysis of the available public data, we projected COVID-19 pandemic peaks and possible ending time and total infected cases in India. Overall, the current pandemic situation with nationwide lockdown is expected to end up by August, 2020, which is a much better scenario than no lockdown in place. Furthermore, we applied our mathematical model to interpret the public data on the total number of infected cases from March, 16, 2020 onwards in two sub regions of India, including Maharastra and Uttar Pradesh, as discussed in the appendix (*D*). With lockdown in place, the peak infection witnessed a significant fall in comparison to the situation if there would have been no lockdown indicating the potential benefit of lockdown in controlling the outspread. Our model suggests a bit relaxed lockdown, followed by an extended period of strict social distancing as one of the most effective control measures to manage COVID-19 spread in days to come.

Further, capturing the impact of the delay on the pandemic transmission, we carried the sensitivity analysis on time delay by varying its values systematically. No qualitative changes have been observed in the infection pattern apart from shifting of the curve resulting in possible domain range for estimated infection.

Owing to significant spatial variations affecting the basic reproduction number *R*_0_, we included spatial heterogeneity by dividing the entire population into smaller sub regions. Based on the technique, *R*_0_ and its time-dependent generalization has been estimated at both regional and sub regional levels. The approach helped us to capture sensible COVID-19 infection dynamics over local and global levels.

The proposed model is an attempt to provide deep insight to analyze the dynamics of COVID-19, helping the involved agencies to arrange and manage crucial resources and design its control strategies. Additionally, the proposed work necessarily made some assumptions when framing the model. We ignore the impact of infection spread due to exposed class [1]. Moreover, the work is based on acquired data for a limited duration of time to fit and estimate the spread of COVID-19. With the release of more epidemic data for India, owing to the regional differences on account of health, education, religion, and per capita income, the key parameters may undergo momentous changes influencing the spread of pandemic among the masses.

## Data Availability

The epidemic bulletin from the World Health Organization provides us with primary data on epidemiological research. We gathered the dataset for COVID-19 in India from the Ministry of Health and Family Welfare (MoHFW), Government of India from March 16, 2020 to April 30, 2020 including the cumulative number of infected cases, the cumulative number of people in recovery and the cumulative number of deaths due to virus. Simultaneously, we collected the state wise total population data for India (projected 2019) from Unique Identification Authority of India (UIDAI), Government of India and migration data to capture the movement of population in different parts of the country from Ministry of Home Affairs, Government of India.

https://www.who.int/emergencies/diseases/novel-coronavirus-2019/events-as-they-happen

https://www.worldometers.info/coronavirus/

https://www.icmr.gov.in/

https://www.mohfw.gov.in/

https://uidai.gov.in/images/state-wise-aadhaar-saturation.pdf

https://censusindia.gov.in/Census_And_You/migrations.aspx/

## V. ACKNOWLEDGMENT

A.K.G. acknowledges financial support from the Science & Engineering Board (SERB), Govt. of India [Grant No.: *CRG*/2019/004669].

## VI. APPENDIX

### A. Epidemiological Model

We consider a large geographic region with total population *N*, partitioned into *n* states labeled by *j* = 1, 2, …*n*. The population within the *j^th^* state is partitioned into susceptibles (*S_j_*), exposures (*E_j_*), infectives (*I_j_*), quarantines (*Q_j_*), recoveries (*R_j_*) and deaths (*D_j_*), where *N_j_ =S_j_* +*E_j_*+*I_j_*+*Q_j_* +*R_j_*+*D_j_* and 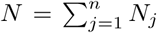. The proposed time delay *SEIQRD* system that captures population and geographic heterogeneities is given by:

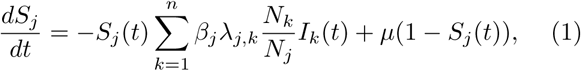

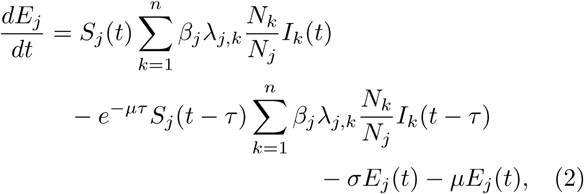

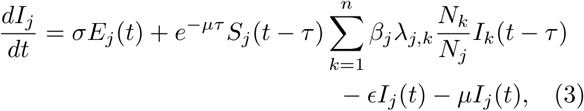

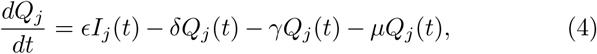

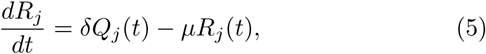

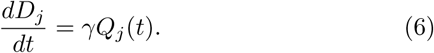

Here *β_j_* > 0 is the average rate of contact between susceptible and infected (people exposed at each time step by infected people) in *j^th^* state; *λ_j,k_* ∈ [0, 1] denotes the population migration factor for the states *j* and *k*. The parameter *σ* ∈ [0, 1] stands for the portion of exposed individuals, which become infectious per time step and *ϵ* ∈ [0, 1] is the quarantine rate from the infection per time step. We call *δ* ∈ [0, 1] the proportion of quarantined people who are cured per time step and *γ* ∈ [0, 1] is the fatality rate due to COVID-19. The parameter *τ* > 0 is the delay time. Further, without affecting the behavior and general aspects of infection, it is assumed that the population in the *j^th^* state is born susceptible with the natural birth rate *μ* > 0 per one individual per time step and the total population remains constant since the natural death rate is considered to be same as the natural birth rate *μ*.

The first term on the R.H.S. of Eq.(1) represents the fragment of the susceptible individual of *j^th^* state who have been exposed to the disease by the infected individuals of the same state and by the infected individuals of the other states, who have moved to the former main state, taking into account the average contact rate; migration coefficients for the states; and the relation between the populations of the states.

Those who were exposed to the infection at time *t* – *τ* and survive the latent period [t – *τ*, *t*] with probability *e^-μτ^* moves to the infected class at time *t* [19, 20]. Moreover, the proportion removed by disease independent mortality for each compartment is given by the last term on R.H.S of each equation. From Eqs.(1) - (6), we get the normality condition *S_j_* (*t*) + *E_j_* (*t*) + *I_j_* (*t*) + *Q_j_* (*t*) + *R_j_* (*t*) + *D_j_* (*t*) = 1, for every state *j* at any time *t*.

### B. Computation of Reproductive number

Following the approaches similar to those taken in [13, 21, 22] with respect to system in Eqs.(1) - (6), it can be verified that the system always has the disease-free equilibrium 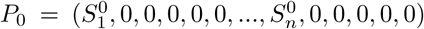 where 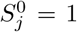 is the equilibrium in the *j^th^* state in the absence of infection, that is *I*_1_ = *I*_2_ = … = *I_n_* = 0. Further, let *R*_0_ = *ρ*(*M*_0_) represents the spectral radius of the matrix M_0_ with entries *m*_0_(*j*, *k*) give by

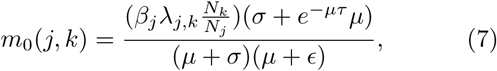

where 1 ≤ *k*, *j* ≤ *n*, then the parameter *R*_0_ is referred to as the basic reproduction number of the considered geographic region. This number is intended to be an indicator of the transmissibility of COVID-19, the outbreak is expected to continue if *R*_0_ > 1 and to end if *R*_0_ < 1. Eqs.(7) shows that the basic reproductive number significantly depends on the average rate of contact between susceptible and infected, population migration factor for the states, state population and time delay. Also, the time-dependent effective basic reproductive number, 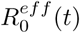, is taken to be 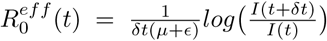. Further, for the *j^th^* state, the basic reproduction number is given by

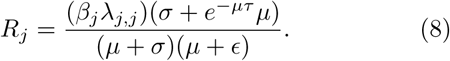

#### C. Model Parameters

The first case of the 2019 – 20 coronavirus pandemic in India was reported on January 30, 2020. We gathered the dataset for COVID-19 in India from the Ministry of Health and Family Welfare (MoHFW), Government of India from March 16, 2020 to April 30, 2020 including the cumulative number of infected cases, the cumulative number of people in recovery and the cumulative number of deaths due to virus [3]. Simultaneously, we collected the state wise total population data for India (projected 2019) from Unique Identification Authority of India (UIDAI), Government of India and migration data to capture the movement of population in different parts of the country from Ministry of Home Affairs, Government of India [23, 24]. State wise population for India is summarized in Table II in order of states as appeared in the map of India in Fig. 1. Therefore, the preliminary estimated parameters that reflect the primary situation of the pandemic in India at the present stage can be summarized as:

**Table II:** State wise population for India (projected 2019) from Unique Identification Authority of India (UIDAI), Government of India [23], in order of states as appeared in map of India in Fig. 1.

| Sr. No. | State | Population |
| --- | --- | --- |
| 1 | Jammu and Kashmir | 13468313 |
| 2 | Ladakh | 279924 |
| 3 | Punjab | 29875481 |
| 4 | Haryana | 27793351 |
| 5 | Uttarakhand | 11140566 |
| 6 | Delhi | 18498192 |
| 7 | Rajasthan | 79584255 |
| 8 | Uttar Pradesh | 233378519 |
| 9 | Himachal Pradesh | 7384022 |
| 10 | Chandigarh | 1142479 |
| 11 | Madhya Pradesh | 83849671 |
| 12 | Bihar | 122256981 |
| 13 | Maharashtra | 121924973 |
| 14 | Karnataka | 66834193 |
| 15 | Telangana | 38919054 |
| 16 | Andhra Pradesh | 53390841 |
| 17 | Goa | 1564349 |
| 18 | Tamil Nadu | 77177540 |
| 19 | Kerala | 35461849 |
| 20 | Chattisgarh | 28989789 |
| 21 | Odisha | 45861035 |
| 22 | West Bengal | 98662146 |
| 23 | Gujrat | 64801901 |
| 24 | Assam | 35080827 |
| 25 | Meghalaya | 3320226 |
| 26 | Jharkhand | 37933898 |
| 27 | Sikkim | 680721 |
| 28 | Arunachal Pradesh | 1548776 |
| 29 | Nagaland | 2218634 |
| 30 | Manipur | 3048861 |
| 31 | Mizoram | 1222134 |
| 32 | Tripura | 4112223 |

### D. Impact on Indian States

We apply our pre-described mathematical model to interpret the public data on the total number of infected cases from March 16, 2020 onwards in sub regions of India, which are published daily by Ministry of Health and Family Welfare (MoHFW), Government of India. Our analysis includes two different regions (states), that is, Maharastra and Uttar Pradesh. It is significant to mention here that we have chosen Maharashtra since it has the highest number of infected cases as of April 30, 2020. Further, to analyze how accurately our model incorporates population heterogeneity, we have chosen Uttar Pradesh since it is the most populated state of India. In Fig. 6(a) and Fig. 6(c), the estimated number of total infected cases is plotted for both the states by solid lines in case of nationwide strict lockdown scenario and actual COVID-19 infection data from March 16, 2020 to April 30, 2020 are marked by circles. Here also the reasonable agreement is observed between actual and forecasted data. We then run the proposed model for chosen parameters for the next 24 months and performed detailed analysis on similar lines for nationwide strict lockdown and no lockdown scenario, as discussed in Section III in Fig. 6(b) and Fig. 6(d). From the numerically obtained outcomes in forward time, with the implementation of lockdown, the peak infection witnessed a significant fall in comparison to the situation if there would have been no lockdown indicating the potential benefit of lockdown in controlling the outspread. Further, as suggested, the situation improves much better if social distancing is enforced strictly along with relaxed lockdown. Using suitably selected parameters, the pandemic transmission patterns can be obtained for any other state of India on similar lines.

**Figure 6:**
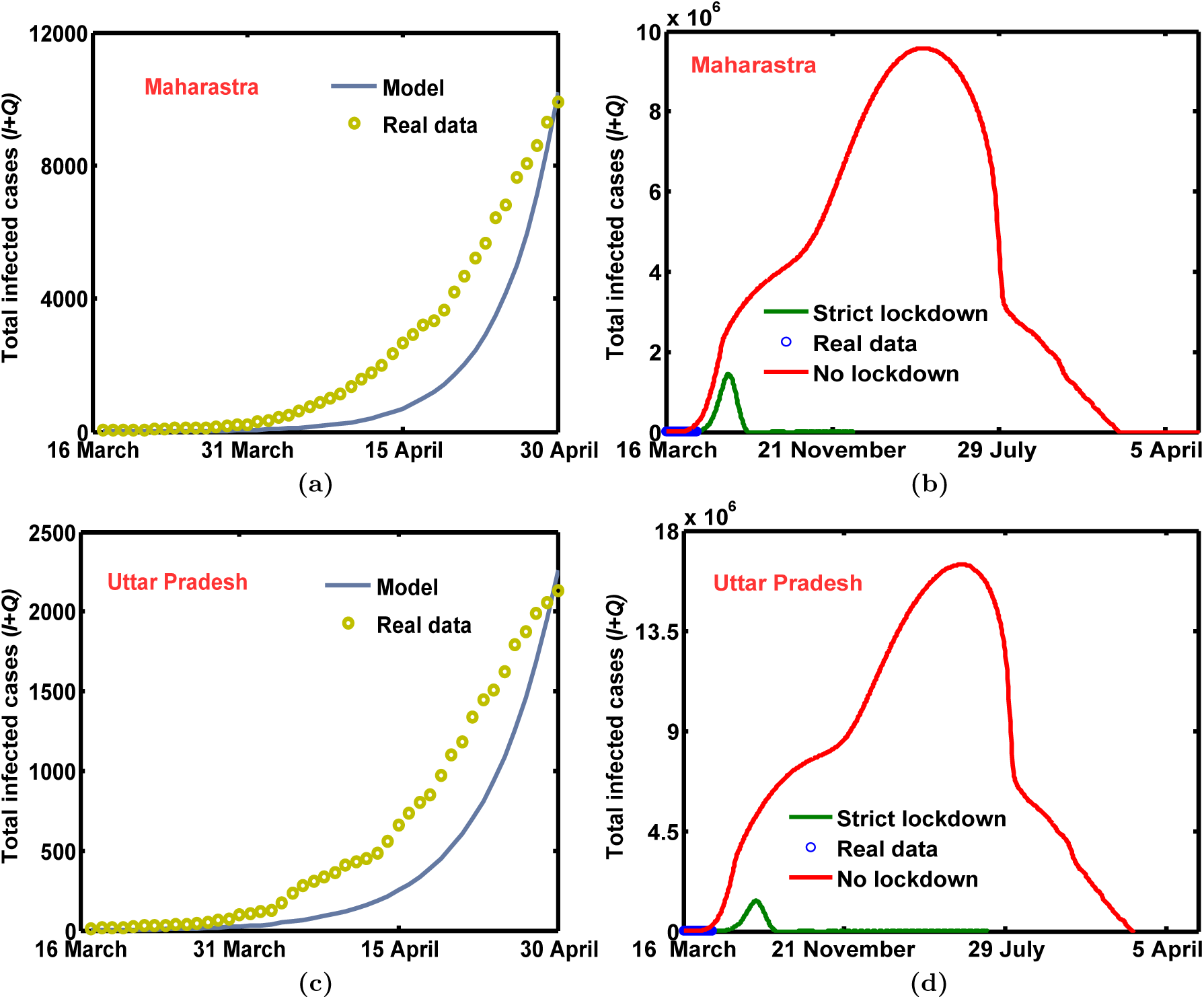
(Color Online) Panels (a) and (c) show fitting of the actual available total COVID-19 infected cases (yellow marker) (*I* + *Q*) for Maharastra and Uttar Pradesh, respectively from 16 March to 30 April, 2020 by the proposed model results (blue). Panels (b) and (d) show total infected cases (*I* + *Q*) in Maharastra, and Uttar Pradesh, respectively with respect to time (in days) in case of strict (green) and no lockdown (red). Blue marker represents the real data. The parameters for (a) and green curve in (b) are: (intrastate population migration factor, interstate population migration factor, *β*, *τ*) = (0.41, 0, 3.4, 7) while in case of no lockdown (red) the chosen values are (0.8, 0.6, 9, 7). In (c) and green curve in (d) these values are (0.393, 0, 3, 8.2) while in case of no lockdown (red) the chosen values are (0.8, 0.6, 9, 8.2). *μ* = 0.0412, *δ* = 0.22, *σ* = 1/7, *ϵ* = 0.85, and *γ* = 0.034 are same in (a-d). In (b) and (d), 29 July, and 5 April belong to 2021, and 2022, respectively.

## Notes

### Competing Interest Statement

The authors have declared no competing interest.

### Funding Statement

A.K.G. acknowledges financial support from the Science and Engineering Board (SERB), Govt. of India [Grant No.: CRG/2019/004669].

